# Pleasure and Peril: The association between sexual risk behavior and sexual pleasure in young adults from the Generation R Study

**DOI:** 10.64898/2026.08.04.26359675

**Authors:** Hella van Stokkom, Linda P. Dekker, Hester Pastoor, Clair A. Enthoven

**Author notes:** **Correspondence to:** Clair A. Enthoven, Department of Child and Adolescent Psychiatry/Psychology, Erasmus MC, Rotterdam, Na Building, Floor 29 |, E. **CRediT author statement: Hella van Stokkom:** Conceptualization, Investigation, Methodology, Visualization, Formal analysis, Writing - Original Draft **Linda P. Dekker:** Conceptualization, Methodology, Supervision, Writing - Review & Editing **Hester Pastoor** Conceptualization, Writing - Review & Editing **Clair A. Enthoven:** Conceptualization, Methodology, Investigation, Supervision, Writing - Review & Editing. **Funding:** The general design of Generation R Study is made possible by financial support from the Erasmus Medical Center and the Erasmus University Rotterdam, the Netherlands Organization for Health Research and Development (ZonMW), the Netherlands Organisation for Scientific Research (NWO), the Ministry of Health, Welfare and Sport and the Ministry of Youth and Families. This study was made possible by the Erasmus Medical Center (MC2 Research Innovation Grant to CAE) and the Erasmus MC Sophia Foundation (“Stichting Vrienden van het Sophia,” Grant WAR25-16 to CAE). The funders had no role in the design and conduct of the study or the writing of the report. **Preregistration:** van Stokkom, Hella, Clair Enthoven, Linda Dekker, and Pauline W. Jansen. Pleasure and Peril: The Association Between Sexual Pleasure and Sexual Risk Behavior in Young Adults. OSF. 2024 Dec 30. doi:10.17605/OSF.IO/DVJEY.

## Abstract

**BACKGROUND:** Sexual pleasure is integral to sexual health, offering important physical and mental benefits. Yet, sex education programs often neglect pleasure, focusing instead on preventing sexual risk behaviors (SRBs), which young adults are particularly vulnerable to.

**AIM:** This study investigates the association between SRBs and sexual pleasure in young adults and whether sex assigned at birth moderates this relationship.

**METHODS:** Embedded within the Generation R cohort, 1010 young adults completed an online questionnaire assessing sexual pleasure using the six subscales of the Amsterdam Sexual Pleasure Inventory (ASPI 1.0) –Arousal Enjoyment, Enjoyment-Related Self-Efficacy, Enjoyment-Related Self-Worth, Interaction Enjoyment, Bonding Enjoyment, and Sexual Experience Enjoyment– and various SRBs including sexual debut <15 years, six or more lifetime partners, frequent unprotected sex, and substance use during sex. Multiple linear regression analyses were performed for each SRB and sexual pleasure subscale, adjusting for demographics, self-esteem, relationship status, socioeconomic status, and psychopathology, with additional stratification by sex assigned at birth.

**OUTCOMES:** The primary outcome measure is sexual pleasure, measured across six domains, examined in relation to SRBs.

**RESULTS:** Fully adjusted regression analyses showed that engaging in SRB was positively associated with several dimensions of sexual pleasure. All SRBs were associated with higher Enjoyment-Related Self-Efficacy (ERSE) scores (p<0.002). Early sexual debut was additionally linked to higher Interaction Enjoyment scores, while having six or more lifetime partners was associated with increased Enjoyment-Related Self-Worth and Sexual Experience Enjoyment scores (p<0.002). Some associations –particularly involving ERSE– were only significant among males. Individuals without partnered sexual experience reported lower sexual pleasure scores.

**CLINICAL IMPLICATIONS:** Incorporating sexual pleasure into sex education could promote a more balanced, realistic understanding of sexuality among young adults, emphasizing both enjoyment and responsible sexual decision-making.

**STRENGHTS & LIMITATIONS:** Key strengths of this study are the use of the multidimensional Amsterdam Sexual Pleasure Inventory (ASPI 1.0) and the large population-based cohort study design, enabling a nuanced and generalizable analysis. This study is limited by potential selection and reporting bias, the cross-sectional design, residual confounding, and the absence of universally agreed-upon thresholds for defining SRBs.

**CONCLUSION:** These findings suggest there is a positive association between engagement in SRB and sexual pleasure, possibly reflecting greater overall sexual experience. The stronger associations observed among males might reflect gendered differences in the role of self-esteem and societal expectations.

## Introduction

Sexual pleasure is an integral part of sexual health [1]. Beyond its intrinsic value, sexual pleasure offers diverse physical and mental health benefits [2], with reduced pleasure strongly predicting sexual distress, even more so than sexual dysfunction [3]. Despite these recognized benefits, sexual pleasure remains largely absent from sex education programs, which predominantly focus on reducing sexual risk behavior (SRB) [4]. SRBs – such as unprotected sex [5,6], sex under the influence of substances [5,6], multiple sexual partners [5,7], and an early sexual debut [5,8] increase the likelihood of adverse outcomes like sexually transmitted infections (STIs) and unintended pregnancy [9]. Adolescents and young adults are particularly vulnerable, as condom use among European youth is declining: from 70% to 61% among males and from 63% to 57% among females between 2014 and 2022 [10]. Although reduced pleasure is often cited by young people as a key reason for not using condoms [11], only 22% of Dutch schools include sexual pleasure in their sex education curricula [12], indicating that sexual pleasure – often overlooked in public health approaches – may be a critical element in young people’s sexual decision-making.

A critical yet underexplored aspect of the relationship between SRBs and sexual pleasure is the role of sex differences. Research on whether males and females experience different levels of sexual pleasure has been mixed [13]. While females and males do not seem to differ in their biological or psychological capacity for pleasure, pleasure gaps found between sexes can be explained by contextual and sociocultural factors [14]. In many societies, heterosexual females’ pleasure is subordinated to males’, and in cultures with greater gender inequality, it is often disregarded or even perceived as dangerous [15]. In these settings, the pleasure gap between sexes is more pronounced [15]. Social norms and cultural expectations also shape how males and females engage in sexual behaviors, including SRB. While young males tend to report higher rates of SRB [6,16], these differences are partly driven by social stigma and safety concerns [14] – females who engage in casual sex face harsher judgment than males, discouraging participation [17]. Notably, when females perceive a safer environment, these differences in casual sex engagement diminish [18].

Research on the relationship between SRBs and sexual pleasure has yielded mixed findings, with some studies finding no association, while others suggest that greater sexual pleasure is linked to lower engagement in SRB (see Supplemental Table S4) [19–28]. Comparisons are complicated by methodological differences and by the frequent use of one-dimensional measures of sexual pleasure focused primarily on the physical sensation or orgasm frequency [19–29]. To address this, Werner et al. [30] proposed a more holistic definition of sexual pleasure, encompassing sensory enjoyment, emotional connection, and confidence in engaging in and deserving pleasure.

Using a newly developed multidimensional measure in a longitudinal, population-based cohort, this study examines (1) whether engagement in SRB is associated with sexual pleasure among young adults, and (2) whether this association is moderated by sex. Understanding these relationships may provide critical insights for developing more effective, pleasure-inclusive sexual health interventions.

## Materials and Methods

### Study design and population

This study was embedded in the Generation R Study, a prospective population-based cohort study from fetal life onwards in Rotterdam, The Netherlands [31]. Pregnant participants living in the study area with a delivery date between 2002 and 2006 were eligible for participation, resulting in 9,778 mothers and their children who enrolled in the study. They were followed up in six waves prenatally and at various ages up to young adulthood.

The current study includes data from an additional online questionnaire distributed to N=3,543 young adults, 18 to 22 years old, who had consented to further research, of whom 1,015 responded (response rate of 29%). While the sexuality data is cross-sectional (from 2024), selected covariates were measured when the participants were nine years old (2011-2015), such as socioeconomic status and mental health symptoms. Participants were included for analyses when they had data on at least one sexual pleasure subscale and on one SRB measurement. This included participants with and without partnered sex experience to ensure diversity within the population, resulting in N=1,010 young adults for the current analyses. For details on the number of included participants with data on each SRB and/or pleasure scale, see Figure 1.

**Figure 1.**
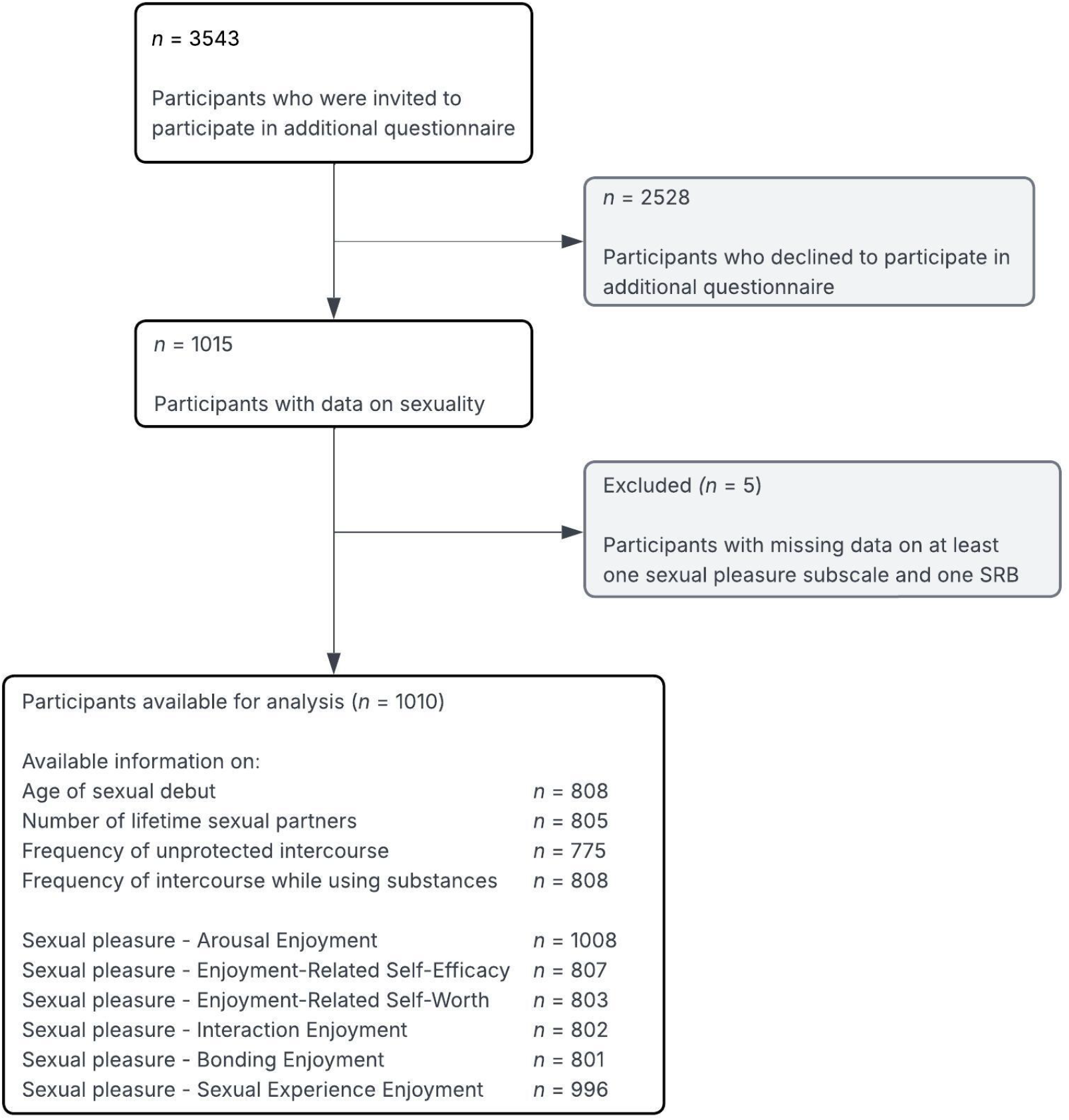
Flowchart of participants included for analysis.

The Medical Ethics Committee of Erasmus MC, University Medical Center Rotterdam, approved the Generation R Study (‘Generation R Phase 4 – Focus on Teens’ Version 5.5. May 2024. METC: 07052024). All participants provided informed consent. Participants were incentivized with a 10-euro voucher upon completing at least 80% of the online questionnaire.

### Measures

#### Sexual risk behavior

Sexual risk behaviors were selected based on previous research [5–8,26]. Participants who reported no partnered sex experience (manual, oral, vaginal, or anal sex; N=201) skipped the SRB questions and were treated as a separate group. Age of sexual debut was categorized into aged >18 years, aged 15 to 18 years and aged <15 years based on the median age of sexual debut in the Netherlands (18.7 years) [11]. Number of lifetime sexual partners were categorized following Pedersen et al. [26] into one, two to five, and six or more. Engaging in sexual intercourse while using substances was categorized into never, sometimes, and often. Engaging in unprotected sexual intercourse (without the intention of becoming pregnant) was categorized into never, sometimes, and often. Participants who responded that they had never had vaginal or anal sex (*n* = 32) were grouped with those reporting no partnered sex experience. Participants who reported having unprotected sex for pregnancy-related reasons were excluded from the analyses for this SRB. A detailed description of the questionnaire items is shown in Supplemental Table S1.

#### Sexual pleasure

Sexual pleasure was measured using the Amsterdam Sexual Pleasure Inventory (ASPI Vol. 1.0), a multidimensional self-report questionnaire that measures sexual pleasure [32]. Only the German version of the questionnaire has undergone validation to date [33]. In its original form, it includes trait and state sections. To reduce respondent burden, this study only used the Trait part of the ASPI, which assesses how one generally experiences sexual pleasure on a 5-point Likert scale. The six Trait scales are: Arousal Enjoyment (5 items; e.g. “I enjoy it when my body reacts to sexual stimuli”), Enjoyment-Related Self-Efficacy (5 items; e.g. “I understand what I need in order to enjoy myself sexually”), Enjoyment-Related Self-Worth (3 items; e.g. “I feel I am worthy of receiving pleasure from my sexpartner”), Interaction Enjoyment (5 items; e.g. “I find It arousing to pleasure my sexpartner during sex”), Bonding Enjoyment (5 items; e.g. “During sex, I enjoy being close to my sexpartner”), and Sexual Experience Enjoyment (4 items; e.g. “I experience sexual pleasure in my life”). Items related to partnered sexual experiences (ASPI items 6-23 and 25) were filtered out for participants without partnered sex experience. Consequently, these individuals (*n* = 201) only had data available on the Arousal Enjoyment and Sexual Experience Enjoyment scales. The available item scores were averaged for each subscale for each respondent, allowing less than 25% of the items per subscale to be missing. Higher scores indicate more sexual pleasure. The Original ASPI Vol. 1.0 can be found at http://dx.doi.org/10.17605/OSF.IO/9HKDE. Most ASPI subscales showed acceptable to excellent internal consistency, except for Enjoyment-Related Self-Worth, which showed questionable internal consistency (α=0.65) (Supplemental Table S2) [34]. A confirmatory factor analysis for all subscales excluding the general Sexual Experience Enjoyment showed overall fit to the data: χ^2^(220) = 1209.73, CFI = .993, TLI = .992, RMSEA = .076, SRMR = .061, with good to strong factor loadings (Supplemental Table S3).

#### Covariates

We selected covariates linked to both SRBs and sexual pleasure based on previous research, including age at assessment (2024, sex assigned at birth [6,13,15–18], sexual orientation [13,35,36], relationship status and duration [13,37,38], socioeconomic status [13,35,39], psychopathology [35,40,41], self-esteem [42,43], and national background [13,44]. A description of the corresponding questionnaire items is shown in Supplemental Table S1.

### Statistical analyses

All analyses were conducted using IBM SPSS Statistics software (version 28.0). Participants’ characteristics were categorized by sex assigned at birth and compared using t-tests for continuous variables and chi-square tests for categorical variables.

To investigate the association between SRBs and sexual pleasure, 24 linear regression models were conducted – one for each of six sexual pleasure domains (dependent variables) and each of four SRBs (independent variables). Covariates were added in a stepwise manner across three models: Model 1 (basic) was unadjusted, Model 2 (partially adjusted) was adjusted for basic demographics (age, sex assigned at birth, sexual orientation, and national background), and Model 3 (main model) was further adjusted for relationship status, psychopathology, socioeconomic status, self-esteem and an interaction term with sex assigned at birth to explore potential moderation. Thus, 24 analyses with three models each were performed. Categorical variables were handled using dummy coding. The least risky behavior served as the reference category for SRBs variables. Stratified analyses were conducted when significant interactions with sex assigned at birth were detected.

Missing values for covariates ranged from 0.2% to 11.6% (relationship status: 0.2%, socioeconomic status: 1.0%, national background: 0.8%, internalizing and externalizing psychopathology scales: 11.6%). To maintain statistical power and reduce potential bias, multiple imputations (m=5) were performed using the Fully Conditional Specification method [45]. To correct for multiple hypothesis testing, a Bonferroni correction [46] was applied, by comparing each p-value with a threshold defined as 0.05 divided by the total number of linear regression analyses – setting the significant threshold at alpha = 0.002 (0.05/24). A post hoc power analysis was conducted using G* Power (version 3.1). All models demonstrated sufficient statistical power to detect the observed effects (power = 1.0). All measures of association are presented as pooled estimates from the imputed datasets, with corresponding 95% confidence intervals (95% CI).

## Results

### Participant characteristics

The participants were on average 20.4 years (SD = 1.0) old, and the majority of them were female (61.6%). Most were European (85.7%), of whom 89.6% were Dutch, identified as heterosexual (77.7%), and had a high socioeconomic status (69.5%). Half of the participants (50.7%) were in a committed relationship, with most relationships lasting over 12 months (54.6%, N=279). Among the 1,010 young adults in the cohort, 201 (19.9%) had no partnered sexual experience. The mean sexual pleasure subscale scores varied between 3.8 and 4.3, and 407 participants had participated in at least one SRB. Table 1 presents the characteristics of the participants, stratified by sex assigned at birth. Females were more likely to identify as bisexual, pansexual, or omnisexual, be in a committed relationship, have partnered sexual experience, have lower internalizing psychopathology scores and scored lower on the sexual pleasure subscales Enjoyment-Related Self-Efficacy and Enjoyment-Related Self-Worth compared to males (p<0.05).

**Table 1.**
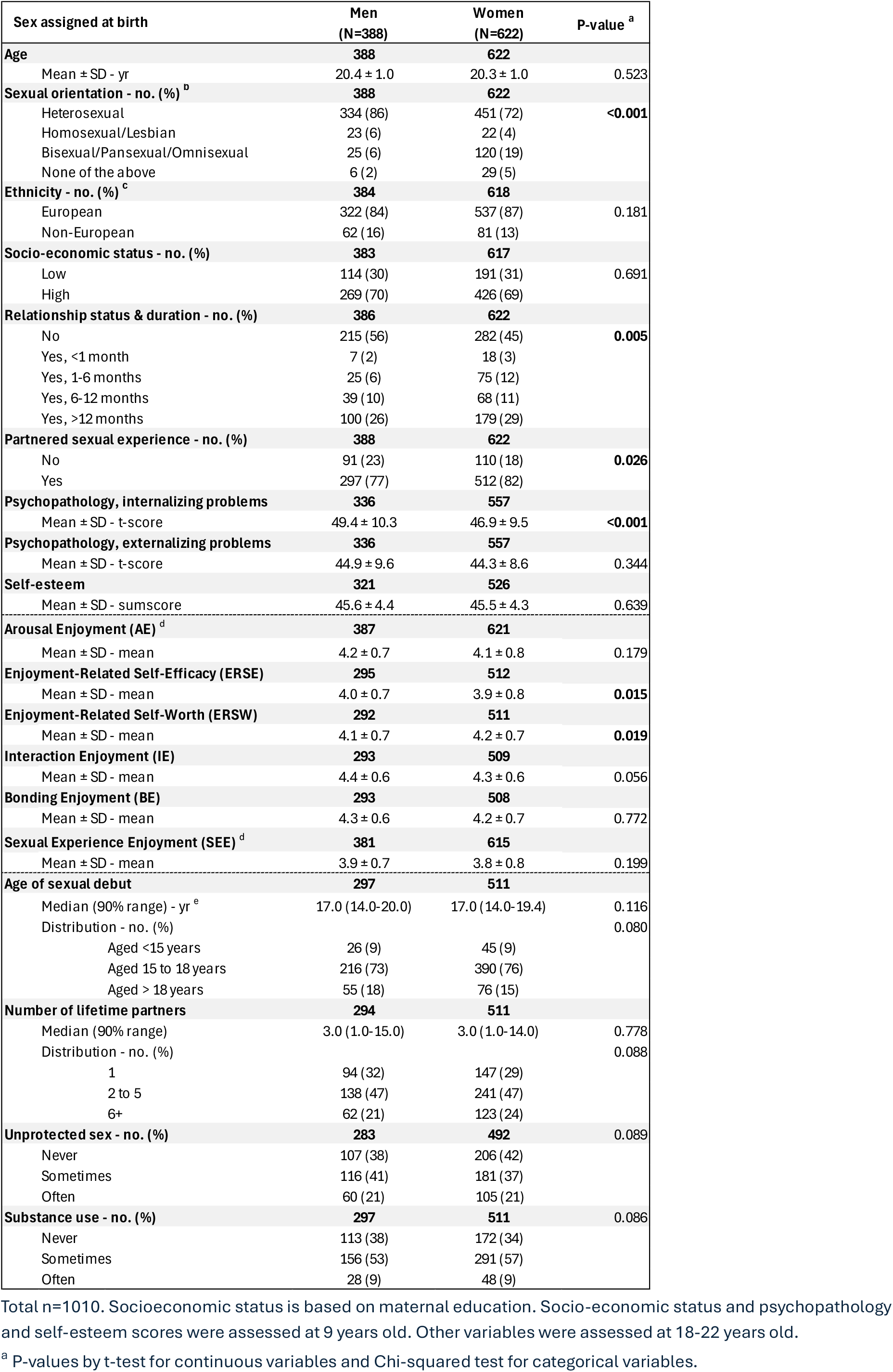
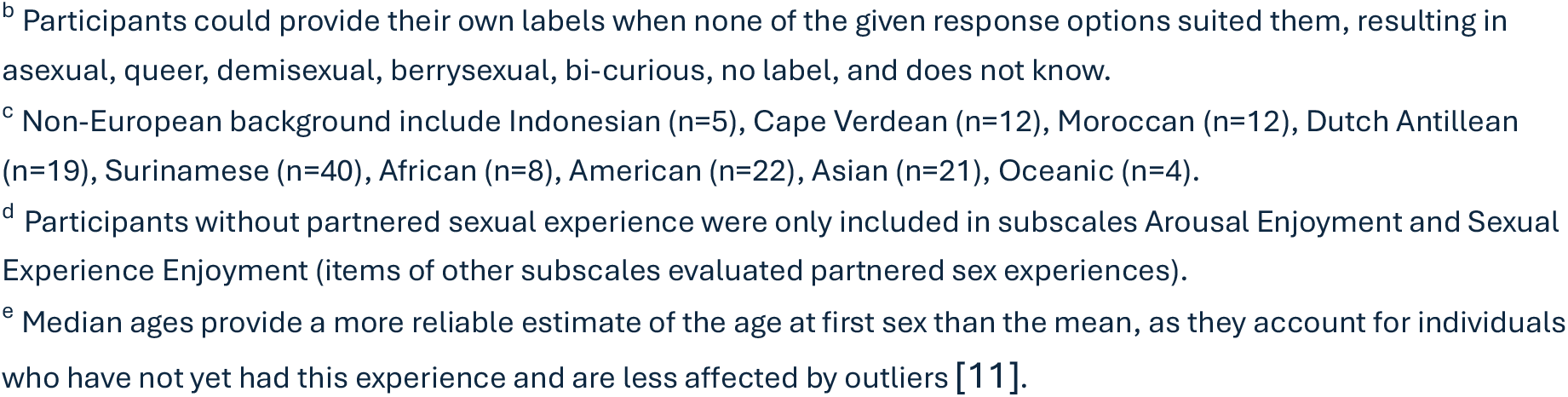
Participant characteristics stratified by sex assigned at birth.

| Sex assigned at birth | Men<br>(N=388) | Women<br>(N=622) | P-value <sup>a</sup> |
| --- | --- | --- | --- |
| **Age** | **388** | **622** |  |
| Mean ± SD - yr | 20.4 ± 1.0 | 20.3 ± 1.0 | 0.523 |
| **Sexual orientation - no. (%) <sup>b</sup>** | **388** | **622** |  |
| Heterosexual | 334 (86) | 451 (72) | <0.001 |
| Homosexual/Lesbian | 23 (6) | 22 (4) |  |
| Bisexual/Pansexual/Omnisexual | 25 (6) | 120 (19) |  |
| None of the above | 6 (2) | 29 (5) |  |
| **Ethnicity - no. (%) <sup>c</sup>** | **384** | **618** |  |
| European | 322 (84) | 537 (87) | 0.181 |
| Non-European | 62 (16) | 81 (13) |  |
| **Socio-economic status - no. (%)** | **383** | **617** |  |
| Low | 114 (30) | 191 (31) | 0.691 |
| High | 269 (70) | 426 (69) |  |
| **Relationship status & duration - no. (%)** | **386** | **622** |  |
| No | 215 (56) | 282 (45) | 0.005 |
| Yes, <1 month | 7 (2) | 18 (3) |  |
| Yes, 1-6 months | 25 (6) | 75 (12) |  |
| Yes, 6-12 months | 39 (10) | 68 (11) |  |
| Yes, >12 months | 100 (26) | 179 (29) |  |
| **Partnered sexual experience - no. (%)** | **388** | **622** |  |
| No | 91 (23) | 110 (18) | 0.026 |
| Yes | 297 (77) | 512 (82) |  |
| **Psychopathology, internalizing problems** | **336** | **557** |  |
| Mean ± SD - t-score | 49.4 ± 10.3 | 46.9 ± 9.5 | <0.001 |
| **Psychopathology, externalizing problems** | **336** | **557** |  |
| Mean ± SD - t-score | 44.9 ± 9.6 | 44.3 ± 8.6 | 0.344 |
| **Self-esteem** | **321** | **526** |  |
| Mean ± SD - sumscore | 45.6 ± 4.4 | 45.5 ± 4.3 | 0.639 |
| **Arousal Enjoyment (AE) <sup>d</sup>** | **387** | **621** |  |
| Mean ± SD - mean | 4.2 ± 0.7 | 4.1 ± 0.8 | 0.179 |
| **Enjoyment-Related Self-Efficacy (ERSE)** | **295** | **512** |  |
| Mean ± SD - mean | 4.0 ± 0.7 | 3.9 ± 0.8 | 0.015 |
| **Enjoyment-Related Self-Worth (ERSW)** | **292** | **511** |  |
| Mean ± SD - mean | 4.1 ± 0.7 | 4.2 ± 0.7 | 0.019 |
| **Interaction Enjoyment (IE)** | **293** | **509** |  |
| Mean ± SD - mean | 4.4 ± 0.6 | 4.3 ± 0.6 | 0.056 |
| **Bonding Enjoyment (BE)** | **293** | **508** |  |
| Mean ± SD - mean | 4.3 ± 0.6 | 4.2 ± 0.7 | 0.772 |
| **Sexual Experience Enjoyment (SEE) <sup>d</sup>** | **381** | **615** |  |
| Mean ± SD - mean | 3.9 ± 0.7 | 3.8 ± 0.8 | 0.199 |
| **Age of sexual debut** | **297** | **511** |  |
| Median (90% range) - yr <sup>e</sup> | 17.0 (14.0-20.0) | 17.0 (14.0-19.4) | 0.116 |
| Distribution - no. (%) |  |  | 0.080 |
| Aged <15 years | 26 (9) | 45 (9) |  |
| Aged 15 to 18 years | 216 (73) | 390 (76) |  |
| Aged > 18 years | 55 (18) | 76 (15) |  |
| **Number of lifetime partners** | **294** | **511** |  |
| Median (90% range) | 3.0 (1.0-15.0) | 3.0 (1.0-14.0) | 0.778 |
| Distribution - no. (%) |  |  | 0.088 |
| 1 | 94 (32) | 147 (29) |  |
| 2 to 5 | 138 (47) | 241 (47) |  |
| 6+ | 62 (21) | 123 (24) |  |
| **Unprotected sex - no. (%)** | **283** | **492** | 0.089 |
| Never | 107 (38) | 206 (42) |  |
| Sometimes | 116 (41) | 181 (37) |  |
| Often | 60 (21) | 105 (21) |  |
| **Substance use - no. (%)** | **297** | **511** | 0.086 |
| Never | 113 (38) | 172 (34) |  |
| Sometimes | 156 (53) | 291 (57) |  |
| Often | 28 (9) | 48 (9) |  |

Total n=1010. Socioeconomic status is based on maternal education. Socio-economic status and psychopathology and self-esteem scores were assessed at 9 years old. Other variables were assessed at 18-22 years old.
<sup>a</sup> P-values by t-test for continuous variables and Chi-squared test for categorical variables.
<sup>b</sup> Participants could provide their own labels when none of the given response options suited them, resulting in asexual, queer, demisexual, berrysexual, bi-curious, no label, and does not know.
<sup>c</sup> Non-European background include Indonesian (n=5), Cape Verdean (n=12), Moroccan (n=12), Dutch Antillean (n=19), Surinamese (n=40), African (n=8), American (n=22), Asian (n=21), Oceanic (n=4).
<sup>d</sup> Participants without partnered sexual experience were only included in subscales Arousal Enjoyment and Sexual Experience Enjoyment (items of other subscales evaluated partnered sex experiences).
<sup>e</sup> Median ages provide a more reliable estimate of the age at first sex than the mean, as they account for individuals who have not yet had this experience and are less affected by outliers [11].

### Associations between Sexual Risk Behavior and Sexual Pleasure

The associations between SRBs and the sexual pleasure subscales from all three models (unadjusted, partially adjusted, and fully adjusted) are presented in Supplemental Table S4. The fully adjusted model showed that after applying multiple testing corrections, having no partnered sexual experience (NPSE) was associated with lower sexual pleasure scores for AE (β: −0.20 to −0.23; 95% CI: −0.34 to −0.08) and SEE (β: −0.18 to −0.25; 95% CI: −0.35 to −0.07) subscales, compared to participants with partnered sexual experience, regardless of SRB category. Having a sexual debut before the age of 15 years was associated with higher ERSE (β: 0.24, 95% CI: 0.11, 0.37) and IE scores (β: 0.24, 95% CI: 0.10, 0.37), compared to having a sexual debut older than 18 years (figure 2a). Having six or more lifetime sexual partners was associated with higher ERSE (β:0.28, 95% CI: 0.15, 0.42), ERSW (β: 0.21, 95% CI: 0.08, 0.35), and SEE (β : 0.23, 95% CI: 0.11, 0.34) scores, compared to having one sexual partner (figure 2b). Often having sex while using substances was associated with higher ERSE scores (β: 0.19, 95% CI: 0.07, 0.30), compared to never having sex while using substances (figure 2c). And often having unprotected sex was associated with higher ERSE scores (β: 0.26, 95% CI: 0.14, 0.38), compared to never having unprotected sex (figure 2d). Some associations were significant in models 1 and 2, but lost significance after further adjusting in model 3 (i.e., sexual intercourse sometimes using substances with AE and often having unprotected sex with SEE). For the Spearman’s correlation matrix between SRBs and sexual pleasure subscales, see Supplemental Figure S1.

**Figure 2.**
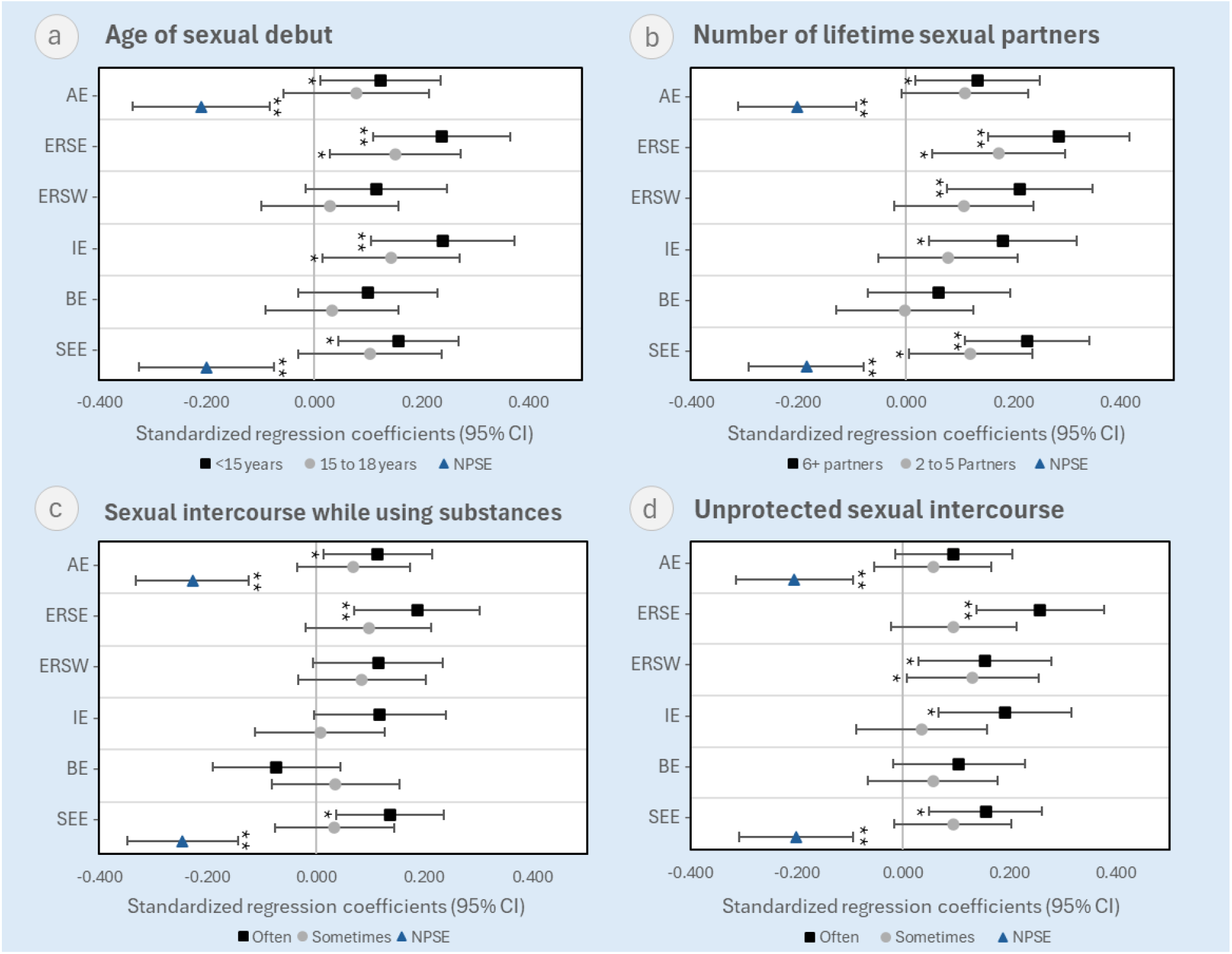
Associations of sexual risk behavior and sexual pleasure in young adults. (a) Age of sexual debut; (b) Number of lifetime sexual partners; (c) Sexual intercourse while using substances; (d) Unprotected sexual intercourse. AE = Arousal Enjoyment. ERSE = Enjoyment-Related Self-Efficacy. ERSW = Enjoyment-Related Self-Worth. IE = Interaction Enjoyment. BE = Bonding Enjoyment. SEE = Sexual Experience Enjoyment. NPSE = No Partnered Sexual Experience. Using standardized regression coefficients *beta*, with their 95% confidence intervals, from model 3 (adjusted for age, sex assigned at birth, sexual orientation, national background, relationship status & duration, socioeconomic status, self-esteem, psychopathology and interaction terms between SRBs and sex assigned at birth). Less risky sexual behaviors were used as a reference. *p-value <0.05. **Bonferroni adjusted p-value <0.002.

### Moderator Effect of Sex

The interaction terms between SRBs and sex assigned at birth were statistically significant for the association between having six or more lifetime partners, often having sex while using substances, and often having unprotected sex with ERSE (p=0.047, p=0.013, p=0.021, respectively), and with ERSW (p=0.025, p=0.006, p=0.044, respectively). Figure 3 illustrates the stronger associations (i.e., steeper slopes) for males compared to females in these relationships. Stratified analyses showed that having six or more lifetime partners and often having unprotected sex were significantly associated with higher ERSE scores in males (figure 3a; β: 0.28, 95% CI: 0.14, 0.42 and figure 3e; β: 0.26, 95% CI: 0.25, 0.66, respectively), but not in females (figure 3a; β: 0.12, 95% CI: 0.02, 0.22 and figure 3e; β: 0.08, 95% CI: −0.01, 0.17, respectively). Sex differences were also observed for the other associations, but these did not remain statistically significant after correcting for multiple testing (figure 3b, 3c and 3f; p<0.05, but not p<0.002). The association between the frequency of sex while using substances and ERSW was not significant for either sex (figure 3d).

**Figure 3.**
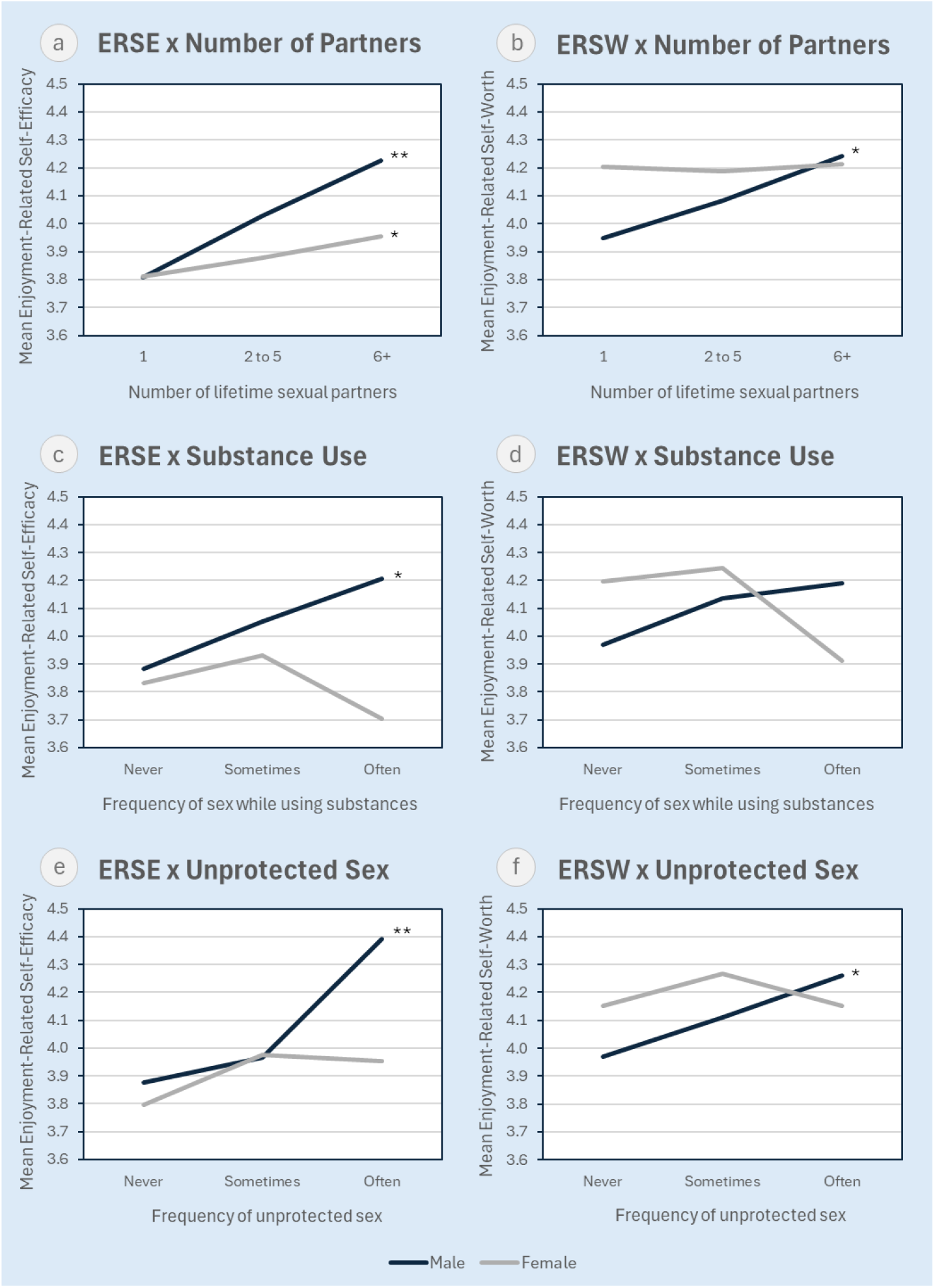
Association between sexual risk behavior and sexual pleasure, stratified by sex. (a) ERSE x Number of Partners; (b) ERSW x Number of Partners; (c) ERSE x Substance Use; (d) ERSW x Substance Use; (e) ERSE x Unprotected Sex; and (f) ERSW x Unprotected Sex. ERSE = Enjoyment-Related Self-Efficacy. ERSW = Enjoyment-Related Self-Worth. The slopes represent the relationship between SRBs and sexual pleasure for males and females. A steeper slope indicates a stronger association. *p<0.05 (fully adjusted model 3). ** Bonferroni adjusted p<0.002 (fully adjusted model 3).

## Discussion

### Observed Associations and Their Implications

Overall, our results showed positive associations between engagement in SRB and sexual pleasure scores, which contradicts much of the existing literature, where studies often report either no association or a negative association (see Supplemental Table S5). However, past studies’ methodology varied significantly, including differences in definitions, measurement tools, and study designs, which complicate direct comparisons. They often used unvalidated, one-dimensional measurement tools, as opposed to the validated, multidimensional ASPI questionnaire. Furthermore, only two out of ten studies in the literature review were conducted in the past five years [24,27], despite a generational shift in young adults’ sexual behavior (in the Netherlands): Condom use is declining, the average age of first sexual encounters is rising, and acceptance of diverse gender identities and sexual orientations is growing [11]. This raises the question of whether this older research is still applicable to this younger generation.

Notably, all four SRBs were associated with significantly higher scores for **Enjoyment-Related Self-Efficacy** (ERSE) compared to participants who did not engage in SRBs. Enjoyment-Related Self-Efficacy reflects the tendency to be confident about engaging in pleasurable sexual activities. One possible explanation is that confidence in sexual encounters may develop into overconfidence, leading individuals to underestimate potential risks and engage in risky behavior.

### Overall Sexual experience

An early sexual debut was significantly associated with higher scores of **Interaction Enjoyment** (IE), and having more than six sexual lifetime partners was associated with significantly higher scores for **Enjoyment-Related Self-Worth** (ERSW) and **Sexual Experience Enjoyment** (SEE). Although these variables are often categorized as SRBs, they may also reflect greater overall sexual experience rather than risk-taking alone. Supporting this interpretation, prior research has shown that individuals with more sexual experience (i.e. a higher number of sexual partners and greater frequency of partnered sex) tend to report higher levels of sexual pleasure [47], likely due to gained sexual self-efficacy and entitlement [48]. Alternatively, individuals who experience greater pleasure from sex may be more inclined to engage in sex more frequently, thereby increasing their opportunities to engage in SRB [47]. Viewed from this perspective, the positive associations observed in the present study may partly reflect the role of overall sexual experience in shaping sexual pleasure rather than the influence of risk behavior itself. This interpretation is further supported by the findings that participants without partnered sexual experience reported lower levels of Arousal Enjoyment (AE) and Sexual Experience Enjoyment (SEE), compared to those with partnered experience.

Several other explanations may also account for the positive associations observed between SRBs and sexual pleasure. For example, experiencing more sexual pleasure may lead to taking more risks. Previous research suggests that heightened sexual arousal may impair judgement and reduce inhibitions, increasing the likelihood of risk-taking during sexual encounters [49]. Conversely, the relationship may operate in the opposite direction, with risk-taking itself enhancing sexual pleasure. For example, reduced pleasure is often cited by young people as a key reason for not using condoms [11] and it has been shown that reduced sexual arousal from condom use influences sexual risk-taking [50].

### Differences between sexes

Differences in pleasure scores between males and females were only observed for Enjoyment-Related Self-Efficacy and Self-Worth, reflecting that men tend to feel more confident in their ability to engage in pleasurable sexual activities and feel more worthy and deserving of positive sexual experiences. However, these differences were minimal (0.1 points), suggesting they may not be clinically significant, which aligns with prior research showing mixed findings in pleasure scores between the sexes [13]. Contrary to earlier studies suggesting that young males engage in SRB more frequently than young females [6,16], no significant sex differences in SRB engagement were observed in this study. However, these findings do align with recent research of a large representative sample of Dutch adolescents and young adults, which reported no difference in the lifetime number of sexual partners and substance use during sex [11].

The association between having six or more lifetime partners and often having unprotected sex with the subscale Enjoyment-Related Self-Efficacy (ERSE) was only significant for males. This may be due to the gender double-standard where social norms encourage women to refrain from having sex and avoid multiple sexual partners, while the peer environment of men is characterized by greater acceptance of sex [51]. Thus, for males, engaging in SRB may reinforce aspects of self-concept tied to masculinity and sexual prowess, in line with societal gender norms [52]. Such reinforcement may translate into higher ERSE scores, even after accounting for global self-esteem. In contrast, for females, engaging in SRB may not provide similar gains in ERSE scores. For females, the pleasure-risk trade-off is likely less favorable, given higher reproductive and health-related consequences [53]. Moreover, sexual self-efficacy in women may be more closely linked to agency and self-advocacy rather than risk-taking. Practicing safer sex often requires asserting personal boundaries and negotiating condom use, behaviors that challenge traditional norms of passive female sexuality [54].

## Strengths and limitations

A key strength of this study is the use of a multidimensional measure of sexual pleasure, the Amsterdam Sexual Pleasure Inventory (ASPI 1.0), enabling a nuanced analysis of its relationship with SRB. Although ASPI scores were left-skewed, this pattern is consistent with the validation study and unlikely to affect regression analyses given the large sample size [33]. The large population-based sample from a well-established cohort allows for a representative analysis and thus the generalizability of the findings. Several limitations should be considered. Selection and reporter bias may have occurred, as individuals more comfortable discussing sexual topics may have been more likely to participate and responses may have been influenced by social desirability, although anonymity may have reduced this risk. Furthermore, the cross-sectional design of this study prevents us from drawing definitive conclusions about causality. Moreover, residual confounding from unmeasured factors such as cultural attitudes, relationship characteristics, and past negative sexual experiences cannot be excluded [13,55]. Additionally, the classification of SRBs remains somewhat arbitrary due to the lack of universally agreed-upon thresholds.

## Implications for sexual health education and interventions

The findings of this study indicate a positive association between SRBs and sexual pleasure, which could initially raise concerns that incorporating sexual pleasure into sex education may encourage risky behavior. However, this relationship likely reflects higher overall sexual experience or activity level – people who have more frequent or varied sexual experiences naturally have increased opportunities for both pleasure and risk-taking. In other words, sexual pleasure and risk-taking are not inherently opposing forces; instead, they’re both linked to higher sexual engagement. Recognizing this, sexual health education could positively emphasize how young people can experience pleasure safely, guiding them to enjoy their sexuality without resorting to risky behaviors using proven approaches such as gain-framed messaging – highlighting the benefits of safer sex – and eroticizing safe sex [56].

Future studies could explore moderating factors such as sexual orientation, cultural differences across diverse populations, and past negative experiences, while using longitudinal and qualitative designs to clarify causal pathways. Additionally, exploring alternative categorizations of SRBs that better capture the spectrum of risk-related behaviors could enhance measurement accuracy and theoretical understanding.

## Conclusion

This study provides important insights into the association between SRBs and sexual pleasure in young adults. Contrary to much of the existing literature, our findings indicate that engagement in SRBs is associated with higher sexual pleasure scores, highlighting the complexity of pleasure and risk in sexual decision-making. Furthermore, certain associations between SRBs and pleasure were significant only for males, suggesting that factors such as societal expectations may differentially influence these experiences. Given the shifting landscape of young adults’ sexual behaviors and attitudes, future research should further explore these associations in diverse populations and consider alternative classifications of risk behaviors. By incorporating sexual pleasure into comprehensive sex education and public health initiatives, we can foster a more balanced and realistic understanding of sexuality – one that promotes both enjoyment and responsible decision-making.

## Data Availability

The data underlying this article cannot be shared publicly because participants of the Generation R study were assured raw data would remain confidential and would not be shared to the public. The data underlying the results presented in the study are available on request with a formal data sharing agreement for researchers who meet the criteria for access to confidential data. Requests should be directed toward the management team of the Generation R study.

## Acknowledgements

We gratefully acknowledge the contribution of the Generation R children and parents, staff, general practitioners, hospitals, midwives, and pharmacies in Rotterdam. We further like to thank Prof. Pauline W. Jansen and Prof. Hanan El Maroun for their feedback on this project and their insights on the general design of the Generation R Study.

## Notes

### Competing Interest Statement

The authors have declared no competing interest.

### Author Declarations

The Medical Ethics Committee of Erasmus MC, University Medical Center Rotterdam gave ethical approval for this work (Generation R Phase 4-Focus on Teens Version 5.5. May 2024. METC: 07052024).

