## Supplementary material for "Pleasure and Peril: The association between sexual risk behavior and sexual pleasure in young adults from the Generation R Study"

**Supplemental Table S1.** Characteristics of variables

| Variable | Question item | Coding |
|---|---|---|
| **Independent variables** | | |
| Age of sexual debut (d) | "How old were you when you did this [consented partnered sex] for the first time?" | (1) No partnered sexual experience<br>(2) Aged >18 years (ref)<br>(3) Aged 15 to 18 years<br>(4) Aged <15 years* |
| Number of lifetime sexual partners (d) | "How many partners have you done this [consented partnered sex] with?" | (1) No partnered sexual experience<br>(2) 1 (ref)<br>(3) 2 to 5<br>(4) 6+* |
| Sexual intercourse while using substances (d) | "Have you ever consumed a lot of alcohol (to the point of being tipsy or drunk) or drugs before or during sex?" | (1) No partnered sexual experience<br>(2) Never (ref)<br>(3) Sometimes<br>(4) Often* |
| Unprotected sexual intercourse (d) [a] | "Have you ever had unprotected sex (vaginal or anal) with someone?"<br>"Did you do this because you were trying to get pregnant or make your partner pregnant?" | (1) No partnered sexual experience<br>(2) Never (ref)<br>(3) Sometimes<br>(4) Often* |
| **Coviarates** | | |
| Age | Age at time of completing questionnaire | N/A [b] |
| Sex | Sex assigned at birth | (0) Male<br>(1) Female |
| Sexual orientation (d) | "How do you see yourself?" | (1) Heterosexual (ref)<br>(2) Homosexual/lesbian<br>(3) Bisexual/pansexual/omnisexual<br>(4) None of the above |
| Relationship status and duration (d) | "Are you currently in a committed relationship?" "When did you start dating each other?" | (1) No (ref)<br>(2) Yes, <1 month<br>(3) Yes, 1-6 months<br>(4) Yes, 6-12 months<br>(5) Yes, >12 months |
| Socioeconomic status | Maternal education (from age-5 wave) | (0) Low (no, primary or secondary education)<br>(1) High (higher than secondary education) |
| Psychopathology | Child Behavior Checklist (CBCL/6-18, school-age version), T-scores, Internalizing and externalizing broadband scales (from age-9 wave) | N/A [b] |
| Self-esteem | Self-Perception Profile for Children (CBSK), sumscores (from age-9 wave) | N/A [b] |
| Ethnicity | Based on country of birth of parents. If the parents were born in different countries, the mother's country of birth determined the ethnic background. | (0) European<br>(1) Non-European |

(d) = dummy variables; (ref) = reference group of dummy variables.

[a] Without the intention of becoming or getting a partner pregnant.

[b] Continuous variable.

*Categories with higher risk behavior.

**Supplemental Table S2.** Reliability ASPI trait subscales

| TRAIT ASPI Subscales | N | Cronbach's alpha |
|---|---|---|
| **Arousal Enjoyment** | 1003 | 0.942 |
| **Enjoyment-Related Self-Efficacy** | 800 | 0.815 |
| **Enjoyment-Related Self-Worth** | 803 | 0.649 |
| **Interaction Enjoyment** | 798 | 0.847 |
| **Bonding Enjoyment** | 792 | 0.883 |
| **Sexual Experience Enjoyment** (all 4 items) | 793 | 0.691 |
| **Sexual Experience Enjoyment** (excluding item about partnered sex) | 992 | 0.668 |

**Supplemental Table S3.** Factor loadings ASPI trait subscales

| Subscale | Items | Factor loadings (95% CI) |
|---|---|---|
| **Arousal enjoyment** | I enjoy it when my body reacts to sexual stimuli | 0.91 (0.90; 0.93) |
| | I love feeling sexual arousal | 0.92 (0.90; 0.94) |
| | I love the sensations of my aroused genitals | 0.88 (0.86; 0.90) |
| | I love it when my erogenous zones are being touched | 0.90 (0.88; 0.92) |
| | I enjoy feeling sexual sensations in my body | 0.95 (0.94; 0.97) |
| **Enjoyment-Related Self-Efficacy** | I know how to shape my sexlife in a way that I really enjoy | 0.86 (0.84; 0.89) |
| | I understand what I need in order to enjoy myself sexually | 0.81 (0.78; 0.84) |
| | I know how to pleasure my sexpartner | 0.73 (0.69; 0.77) |
| | I can engage in partnersex in a way that I really enjoy | 0.91 (0.89; 0.93) |
| | I can masturbate in a way that I really enjoy | 0.52 (0.46; 0.58) |
| **Enjoyment-Related Self-Worth** | I feel I am worthy of receiving plesaure from my sexpartner | 0.89 (0.84; 0.95) |
| | During partnersex, I neglect my own pleasure | 0.60 (0.54; 0.65) |
| | My sexual pleasure is irrelevant | 0.66 (0.61; 0.72) |
| **Interaction Enjoyment** | I find it arousing to entice my sexpartner into having sex | 0.72 (0.67; 0.76) |
| | I feel fulfilled when my sexpartner enjoys themselves during sex | 0.89 (0.87; 0.92) |
| | I find it arousing to pleasure my sexpartner during sex | 0.91 (0.89; 0.93) |
| | I enjoy stimulating my sexpartner during sex | 0.89 (0.87; 0.91) |
| | I enjoy it when my sexpartner stimulates me during sex | 0.88 (0.85; 0.92) |
| **Bonding Enjoyment** | During sex, I enjoy being close to my sexpartner | 0.91 (0.88; 0.94) |
| | During sex, I feel connected to my sexpartner | 0.85 (0.82; 0.88) |
| | During sex, I enjoy the affection between me and my sexpartner | 0.91 (0.89; 0.94) |
| | During partnersex, I enjoy the feeling of security | 0.79 (0.76; 0.82) |
| | Sex brings me closer to my sexpartner | 0.79 (0.76; 0.83) |

CI= Confidence Interval; Confirmatory Factor Analyses was conducted among participants with partnered sexual experience.

**Supplemental Table S4.** Associations of sexual risk behavior and sexual pleasure in young adults.

| | Sexual Pleasure Subscales | | | | | | | | | | | |
|---|---|---|---|---|---|---|---|---|---|---|---|---|
| | AE | | ERSE | | ERSW | | IE | | BE | | SEE | |
| | Unstandardized B (95% CI) | Standardized β (95% CI) | Unstandardized B (95% CI) | Standardized β (95% CI) | Unstandardized B (95% CI) | Standardized β (95% CI) | Unstandardized B (95% CI) | Standardized β (95% CI) | Unstandardized B (95% CI) | Standardized β (95% CI) | Unstandardized B (95% CI) | Standardized β (95% CI) |
| **Age of sexual debut** | *n* = 1007 | | *n* = 806 | | *n* = 802 | | *n* = 801 | | *n* = 800 | | *n* = 995 | |
| Model 1 (Constant) | 4.27 | | 3.72 | | 4.07 | | 4.25 | | 4.21 | | 3.90 | |
| NPSE | -0.69 (-0.84, -0.53)** | -0.36 (-0.45, -0.28) | N/A | | N/A | | N/A | | N/A | | -0.67 (-0.83, -0.52)** | -0.35 (-0.43, -0.26) |
| >18 years | *Reference* | | *Reference* | | *Reference* | | *Reference* | | *Reference* | | *Reference* | |
| 15-18 years | 0.02 (-0.11, 0.16) | 0.01 (-0.07, 0.10) | 0.21 (0.08, 0.35)* | 0.13 (0.04, 0.21) | 0.07 (-0.06, 0.21) | 0.05 (-0.04, 0.13) | 0.08 (-0.03, 0.19) | 0.06 (-0.02, 0.14) | 0.04 (-0.09, 0.16) | 0.02 (-0.06, 0.11) | 0.08 (-0.06, 0.21) | 0.05 (-0.04, 0.13) |
| <15 years | 0.13 (-0.07, 0.33) | 0.04 (-0.02, 0.11) | 0.50 (0.29, 0.71)** | 0.19 (0.11, 0.28) | 0.26 (0.06, 0.47)* | 0.11 (0.03, 0.19) | 0.27 (0.10, 0.44)** | 0.13 (0.05, 0.21) | 0.22 (0.03, 0.41)* | 0.09 (0.01, 0.18) | 0.26 (0.05, 0.47)* | 0.09 (0.02, 0.16) |
| Model 2 (Constant) | 3.46 | | 2.16 | | 3.42 | | 2.68 | | 3.02 | | 2.67 | |
| NPSE | -0.64 (-0.80, -0.49)** | -0.34 (-0.43, -0.26) | N/A | | N/A | | N/A | | N/A | | -0.62 (-0.78, -0.46)** | -0.32 (-0.40, -0.23) |
| >18 years | *Reference* | | *Reference* | | *Reference* | | *Reference* | | *Reference* | | *Reference* | |
| 15-18 years | 0.05 (-0.08, 0.18) | 0.03 (-0.06, 0.12) | 0.26 (0.12, 0.40)** | 0.15 (0.07, 0.24) | 0.07 (-0.07, 0.20) | 0.04 (-0.04, 0.12) | 0.12 (0.01, 0.23)* | 0.09 (0.01, 0.17) | 0.06 (-0.07, 0.19) | 0.04 (-0.04, 0.12) | 0.12 (-0.01, 0.26) | 0.08 (-0.01, 0.16) |
| <15 years | 0.15 (-0.05, 0.35) | 0.05 (-0.02, 0.12) | 0.54 (0.33, 0.75)** | 0.21 (0.12, 0.29) | 0.25 (0.05, 0.45)* | 0.10 (0.02, 0.19) | 0.31 (0.14, 0.48)** | 0.15 (0.07, 0.23) | 0.23 (0.04, 0.42)* | 0.10 (0.01, 0.18) | 0.30 (0.09, 0.51)* | 0.10 (0.03, 0.17) |
| Model 3 (Constant) | 3.13 | | 1.76 | | 3.31 | | 2.41 | | 3.10 | | 2.16 | |
| NPSE | -0.40 (-0.64, -0.16)** | -0.21 (-0.34, -0.08) | N/A | | N/A | | N/A | | N/A | | -0.39 (-0.63, -0.15)** | -0.20 (-0.33, -0.07) |
| >18 years | *Reference* | | *Reference* | | *Reference* | | *Reference* | | *Reference* | | *Reference* | |
| 15-18 years | 0.12 (-0.09, 0.33) | 0.08 (-0.06, 0.22) | 0.26 (0.05, 0.46)* | 0.15 (0.03, 0.28) | 0.05 (-0.15, 0.25) | 0.03 (-0.10, 0.16) | 0.19 (0.02, 0.36)* | 0.14 (0.01, 0.27) | 0.05 (-0.14, 0.24) | 0.03 (-0.09, 0.16) | 0.17 (-0.04, 0.38) | 0.11 (-0.03, 0.24) |
| <15 years | 0.36 (0.04, 0.69)* | 0.12 (0.01, 0.24) | 0.62 (0.29, 0.95)** | 0.24 (0.11, 0.37) | 0.28 (-0.04, 0.61) | 0.12 (-0.02, 0.25) | 0.50 (0.22, 0.77)** | 0.24 (0.10, 0.37) | 0.24 (-0.07, 0.54) | 0.10 (-0.03, 0.23) | 0.48 (0.14, 0.82)* | 0.16 (0.05, 0.27) |
| **Number of lifetime partners** | *n* = 1004 | | *n* = 804 | | *n* = 800 | | *n* = 799 | | *n* = 798 | | *n* = 993 | |
| Model 1 (Constant) | 4.25 | | 3.81 | | 4.11 | | 4.28 | | 4.28 | | 3.88 | |
| NPSE | -0.66 (-0.79, -0.53)** | -0.35 (-0.42, -0.28) | N/A | | N/A | | N/A | | N/A | | -0.66 (-0.79, -0.52)** | -0.34 (-0.41, -0.27) |
| 1 | *Reference* | | *Reference* | | *Reference* | | *Reference* | | *Reference* | | *Reference* | |
| 2 to 5 | 0.04 (-0.07, 0.15) | 0.03 (-0.05, 0.10) | 0.12 (0.005, 0.24)* | 0.08 (0.003, 0.16) | 0.04 (-0.07, 0.16) | 0.03 (-0.05, 0.11) | 0.05 (-0.05, 0.14) | 0.04 (-0.04, 0.12) | -0.05 (-0.16, 0.06) | -0.04 (-0.12, 0.04) | 0.07 (-0.05, 0.18) | 0.04 (-0.03, 0.12) |
| 6+ | 0.14 (0.01, 0.28)* | 0.07 (0.005, 0.14) | 0.24 (0.10, 0.38)** | 0.14 (0.05, 0.22) | 0.12 (-0.01, 0.25) | 0.07 (-0.01, 0.15) | 0.15 (0.03, 0.26)* | 0.11 (0.02, 0.19) | -0.004 (-0.13, 0.12) | -0.003 (-0.08, 0.08) | 0.28 (0.14, 0.42)** | 0.14 (0.07, 0.21) |
| Model 2 (Constant) | 3.75 | | 3.08 | | 3.73 | | 3.18 | | 3.16 | | 3.24 | |
| NPSE | -0.65 (-0.78, -0.52)** | -0.35 (-0.42, -0.28) | N/A | | N/A | | N/A | | N/A | | -0.65 (-0.78, -0.52)** | -0.33 (-0.40, -0.26) |
| 1 | *Reference* | | *Reference* | | *Reference* | | *Reference* | | *Reference* | | *Reference* | |
| 2 to 5 | 0.04 (-0.08, 0.15) | 0.02 (-0.05, 0.10) | 0.12 (-0.003, 0.23) | 0.08 (-0.003, 0.16) | 0.04 (-0.08, 0.15) | 0.03 (-0.05, 0.11) | 0.04 (-0.06, 0.13) | 0.03 (-0.05, 0.11) | -0.06 (-0.17, 0.05) | -0.04 (-0.13, 0.04) | 0.06 (-0.05, 0.18) | 0.04 (-0.03, 0.11) |
| 6+ | 0.14 (0.002, 0.27)* | 0.07 (0.0001, 0.14) | 0.22 (0.08, 0.36)* | 0.13 (0.04, 0.21) | 0.10 (-0.04, 0.23) | 0.06 (-0.02, 0.14) | 0.12 (0.004, 0.23)* | 0.08 (0.002, 0.17) | -0.04 (-0.17, 0.09) | -0.03 (-0.11, 0.06) | 0.27 (0.13, 0.41)** | 0.13 (0.06, 0.20) |
| Model 3 (Constant) | 3.49 | | 2.81 | | 3.62 | | 3.10 | | 3.28 | | 2.81 | |
| NPSE | -0.38 (-0.59, -0.17)** | -0.20 (-0.31, -0.09) | N/A | | N/A | | N/A | | N/A | | -0.36 (-0.57, -0.15)** | -0.18 (-0.29, -0.08) |
| 1 | *Reference* | | *Reference* | | *Reference* | | *Reference* | | *Reference* | | *Reference* | |
| 2 to 5 | 0.17 (-0.01, 0.35) | 0.11 (-0.01, 0.23) | 0.25 (0.07, 0.44)* | 0.17 (0.05, 0.30) | 0.15 (-0.03, 0.33) | 0.11 (-0.02, 0.24) | 0.09 (-0.06, 0.25) | 0.08 (-0.05, 0.21) | -0.003 (-0.17, 0.16) | -0.002 (-0.13, 0.13) | 0.19 (0.01, 0.38)* | 0.12 (0.003, 0.24) |
| 6+ | 0.26 (0.03, 0.49)* | 0.13 (0.02, 0.25) | 0.50 (0.27, 0.72)** | 0.28 (0.15, 0.42) | 0.35 (0.13, 0.57)** | 0.21 (0.08, 0.35) | 0.25 (0.06, 0.44)* | 0.18 (0.04, 0.32) | 0.10 (-0.11, 0.30) | 0.06 (-0.07, 0.20) | 0.45 (0.22, 0.68)** | 0.23 (0.11, 0.34) |

| | Sexual Pleasure Subscales | | | | | | | | | | | |
|---|---|---|---|---|---|---|---|---|---|---|---|---|
| | AE | | ERSE | | ERSW | | IE | | BE | | SEE | |
| | Unstandardized B (95% CI) | Standardized β (95% CI) | Unstandardized B (95% CI) | Standardized β (95% CI) | Unstandardized B (95% CI) | Standardized β (95% CI) | Unstandardized B (95% CI) | Standardized β (95% CI) | Unstandardized B (95% CI) | Standardized β (95% CI) | Unstandardized B (95% CI) | Standardized β (95% CI) |
| **Sex while using substances** | *n* = 1007 | | *n* = 806 | | *n* = 802 | | *n* = 801 | | *n* = 800 | | *n* = 995 | |
| Model 1 (Constant) | 4.17 | | 3.85 | | 4.11 | | 4.27 | | 4.21 | | 3.87 | |
| NPSE | **-0.59 (-0.71, -0.46)**** | **-0.31 (-0.38, -0.24)** | N/A | | N/A | | N/A | | N/A | | **-0.64 (-0.77, -0.51)**** | **-0.33 (-0.40, -0.26)** |
| Never | *Reference* | | *Reference* | | *Reference* | | *Reference* | | *Reference* | | *Reference* | |
| Sometimes | **0.19 (0.09, 0.30)**** | **0.13 (0.06, 0.20)** | 0.12 (0.01, 0.23)* | 0.08 (0.01, 0.16) | 0.10 (-0.01, 0.20) | 0.07 (-0.01, 0.15) | 0.10 (0.01, 0.18)* | 0.08 (0.01, 0.16) | 0.11 (0.01, 0.21)* | 0.08 (0.01, 0.16) | 0.16 (0.05, 0.26)* | 0.10 (0.03, 0.17) |
| Often | 0.24 (0.06, 0.41)* | 0.08 (0.02, 0.15) | 0.04 (-0.15, 0.22) | 0.01 (-0.06, 0.09) | -0.10 (-0.27, 0.08) | -0.04 (-0.12, 0.03) | 0.17 (0.02, 0.31)* | 0.08 (0.01, 0.16) | -0.15 (-0.32, 0.01) | -0.07 (-0.14, 0.01) | 0.27 (0.09, 0.46)* | 0.09 (0.03, 0.16) |
| Model 2 (Constant) | 3.56 | | 2.80 | | 3.59 | | 3.01 | | 3.21 | | 2.94 | |
| NPSE | **-0.57 (-0.70, -0.45)**** | **-0.30 (-0.37, -0.24)** | N/A | | N/A | | N/A | | N/A | | **-0.63 (-0.76, -0.50)**** | **-0.32 (-0.39, -0.26)** |
| Never | *Reference* | | *Reference* | | *Reference* | | *Reference* | | *Reference* | | *Reference* | |
| Sometimes | **0.18 (0.08, 0.29)**** | **0.12 (0.05, 0.19)** | 0.12 (0.01, 0.22)* | 0.08 (0.003, 0.15) | 0.08 (-0.02, 0.18) | 0.06 (-0.02, 0.13) | 0.09 (0.003, 0.18)* | 0.08 (0.002, 0.15) | 0.10 (0.002, 0.20)* | 0.07 (0.002, 0.15) | 0.15 (0.04, 0.26)* | 0.10 (0.03, 0.17) |
| Often | 0.23 (0.06, 0.40)* | 0.08 (0.02, 0.14) | 0.03 (-0.15, 0.22) | 0.01 (-0.06, 0.09) | -0.11 (-0.28, 0.06) | -0.05 (-0.12, 0.03) | 0.16 (0.01, 0.31)* | 0.08 (0.004, 0.15) | -0.16 (-0.33, 0.004) | -0.07 (-0.15, 0.003) | 0.27 (0.09, 0.45)* | 0.09 (0.03, 0.15) |
| Model 3 (Constant) | 3.43 | | 2.45 | | 3.42 | | 2.96 | | 3.33 | | 2.62 | |
| NPSE | **-0.43 (-0.62, -0.23)**** | **-0.23 (-0.33, -0.12)** | N/A | | N/A | | N/A | | N/A | | **-0.48 (-0.68, -0.28)**** | **-0.25 (-0.35, -0.14)** |
| Never | *Reference* | | *Reference* | | *Reference* | | *Reference* | | *Reference* | | *Reference* | |
| Sometimes | 0.10 (-0.06, 0.27) | 0.07 (-0.04, 0.18) | 0.14 (-0.03, 0.31) | 0.10 (-0.02, 0.21) | 0.12 (-0.05, 0.28) | 0.08 (-0.03, 0.20) | 0.01 (-0.13, 0.15) | 0.01 (-0.11, 0.13) | 0.05 (-0.11, 0.20) | 0.04 (-0.08, 0.15) | 0.05 (-0.12, 0.23) | 0.03 (-0.08, 0.15) |
| Often | 0.32 (0.04, 0.61)* | 0.11 (0.01, 0.22) | **0.47 (0.18, 0.76)**** | **0.19 (0.07, 0.30)** | 0.27 (-0.01, 0.55) | 0.11 (-0.01, 0.24) | 0.24 (-0.01, 0.48) | 0.12 (-0.004, 0.24) | -0.16 (-0.43, 0.10) | -0.07 (-0.19, 0.05) | 0.40 (0.11, 0.70)* | 0.14 (0.03, 0.24) |
| **Unprotected sex** | *n* = 1006 | | *n* = 805 | | *n* = 801 | | *n* = 800 | | *n* = 799 | | *n* = 994 | |
| Model 1 (Constant) | 4.27 | | 3.82 | | 4.09 | | 4.28 | | 4.22 | | 3.89 | |
| NPSE | **-0.59 (-0.71, -0.47)**** | **-0.33 (-0.40, -0.27)** | -0.40 (-0.67, -0.14)* | **-0.11 (-0.18, -0.04)** | -0.03 (-0.28, 0.23) | -0.01 (-0.08, 0.06) | 0.06 (-0.16, 0.28) | 0.02 (-0.05, 0.09) | 0.04 (-0.21, 0.28) | 0.01 (-0.06, 0.08) | **-0.60 (-0.72, -0.47)**** | **-0.32 (-0.39, -0.25)** |
| Never | *Reference* | | *Reference* | | *Reference* | | *Reference* | | *Reference* | | *Reference* | |
| Sometimes | 0.02 (-0.09, 0.13) | 0.01 (-0.06, 0.08) | 0.15 (0.04, 0.26)* | 0.10 (0.02, 0.17) | 0.12 (0.01, 0.23)* | 0.08 (0.003, 0.16) | 0.06 (-0.04, 0.15) | 0.05 (-0.03, 0.12) | 0.05 (-0.05, 0.16) | 0.04 (-0.04, 0.12) | 0.12 (0.005, 0.23)* | 0.07 (0.002, 0.14) |
| Often | 0.12 (-0.01, 0.25) | 0.06 (-0.01, 0.13) | **0.29 (0.15, 0.42)**** | **0.16 (0.08, 0.23)** | 0.10 (-0.03, 0.23) | 0.06 (-0.02, 0.14) | 0.17 (0.06, 0.28)* | 0.12 (0.04, 0.20) | 0.08 (-0.05, 0.20) | 0.05 (-0.03, 0.13) | **0.23 (0.10, 0.37)**** | **0.11 (0.05, 0.18)** |
| Model 2 (Constant) | 3.52 | | 2.89 | | 3.63 | | 3.03 | | 3.23 | | 2.91 | |
| NPSE | **-0.58 (-0.70, -0.46)**** | **-0.33 (-0.39, -0.26)** | -0.42 (-0.68, -0.15)* | -0.11 (-0.18, -0.04) | 0.02 (-0.24, 0.27) | 0.01 (-0.07, 0.08) | 0.05 (-0.18, 0.27) | 0.01 (-0.06, 0.09) | 0.03 (-0.22, 0.27) | 0.01 (-0.07, 0.08) | **-0.59 (-0.72, -0.47)**** | **-0.32 (-0.39, -0.25)** |
| Never | *Reference* | | *Reference* | | *Reference* | | *Reference* | | *Reference* | | *Reference* | |
| Sometimes | 0.02 (-0.09, 0.13) | 0.01 (-0.06, 0.08) | 0.14 (0.03, 0.25)* | 0.09 (0.02, 0.17) | 0.12 (0.01, 0.23)* | 0.09 (0.01, 0.16) | 0.05 (-0.04, 0.14) | 0.04 (-0.03, 0.12) | 0.05 (-0.06, 0.15) | 0.04 (-0.04, 0.12) | 0.12 (0.01, 0.23)* | 0.07 (0.002, 0.14) |
| Often | 0.12 (-0.01, 0.25) | 0.06 (-0.01, 0.13) | **0.28 (0.14, 0.41)**** | **0.15 (0.08, 0.23)** | 0.10 (-0.03, 0.23) | 0.06 (-0.02, 0.13) | 0.17 (0.06, 0.28)* | 0.12 (0.04, 0.19) | 0.07 (-0.05, 0.19) | 0.04 (-0.03, 0.12) | **0.23 (0.10, 0.37)**** | **0.11 (0.04, 0.18)** |
| Model 3 (Constant) | 3.22 | | 2.44 | | 3.42 | | 2.82 | | 3.23 | | 2.45 | |
| NPSE | **-0.36 (-0.56, -0.17)**** | **-0.20 (-0.31, -0.09)** | -0.28 (-0.69, 0.13) | -0.07 (-0.18, 0.03) | -0.09 (-0.49, 0.30) | -0.03 (-0.14, 0.09) | -0.04 (-0.39, 0.32) | -0.01 (-0.13, 0.10) | 0.23 (-0.14, 0.61) | 0.07 (-0.04, 0.18) | **-0.37 (-0.57, -0.18)**** | **-0.20 (-0.31, -0.09)** |
| Never | *Reference* | | *Reference* | | *Reference* | | *Reference* | | *Reference* | | *Reference* | |
| Sometimes | 0.09 (-0.09, 0.28) | 0.06 (-0.06, 0.17) | 0.15 (-0.03, 0.32) | 0.10 (-0.02, 0.21) | 0.19 (0.01, 0.36)* | 0.13 (0.01, 0.26) | 0.04 (-0.11, 0.19) | 0.04 (-0.09, 0.16) | 0.08 (-0.09, 0.24) | 0.06 (-0.07, 0.18) | 0.16 (-0.02, 0.35) | 0.09 (-0.02, 0.21) |
| Often | 0.19 (-0.03, 0.41) | 0.10 (-0.02, 0.21) | **0.47 (0.25, 0.68)**** | **0.26 (0.14, 0.38)** | 0.26 (0.05, 0.48)* | 0.15 (0.03, 0.28) | 0.28 (0.10, 0.46)* | 0.19 (0.06, 0.32) | 0.17 (-0.03, 0.37) | 0.10 (-0.02, 0.23) | 0.33 (0.10, 0.55)* | 0.15 (0.05, 0.26) |

### Supplementary Table S4. (continued)

NPSE = No Partnered Sexual Experience. AE = Arousal Enjoyment. ERSE = Enjoyment-Related Self-Efficacy. ERSW = Enjoyment-Related Self-Worth. IE = Interaction Enjoyment. BE = Bonding Enjoyment. SEE = Sexual Experience Enjoyment. 'n' represents the number of participants used for the analysis. It shows unstandardized regression coefficients (B) with their 95% Confidence Intervals and standardized regression coefficients (β), derived from linear regression models (as pooled estimates from the imputed datasets). Model 1 is unadjusted. Model 2 is adjusted for age, sex assigned at birth, national background, and sexual orientation. Model 3 (main model) is additionally adjusted for self-esteem, relationship status & duration, socioeconomic status, and psychopathology, including interaction terms for sex assigned at birth. Less risky sexual behaviors were used as a reference. *p-value <0.05. **p-value <0.002 (after multiple testing correction).

**Supplementary Table S5**. Characteristics and findings of studies on the association between sexual risk behaviors and sexual satisfaction.

| Author | N | Country | Age in years | Statistics | Measure of sexual pleasure | Findings | |
|---|---|---|---|---|---|---|---|
| Auslander et al 2007 | 313 | USA | M = 19.7 (Adolescent/Young Adult) | Bivariate logistic regression | 7-item scale (sexual pleasure and sexual excitement in current relationship) | • Number of lifetime sexual partners<br>• % of condomprotected vaginal sexual episodes in last 3 months | –<br>– |
| Haavio-Mannila & Kontula 1997 | 2250 | Finland | Range 18-54 | Stepwise multiple regression | 1-item ("pleasurableness of sexual intercourse") | • Early sexual debut | ○ |
| Heiman et al 2011 | 400 | Brazil, Germany, Japan, Spain, USA | Median = 53.5 (Middle-aged/Older couples) | Logit models | Single item (sexual satisfaction past 4 weeks) | • Number of lifetime sexual partners | – *(only for men)* |
| Higgins et al 2008 | 189 | USA | M = 24.5 (Women only) | Multiple linear regression | 3-item scale | • Condom use *(compared to other contraceptive methods)* | ○ |
| Higgins et al 2011 | 2168 | USA | M = 20.2 | Multivariate logistic regression | 2-item scale (physical and psychological sexual satisfaction) | • Contraception use<br>• Number of partners in last year<br>• Perceived likelihood of contracting a STI | ○<br>○<br>○ |
| Klein et al 2022 | 3472 | Germany | Range 18-75 | Logistic regression | Amsterdam Sexual Pleasure Index (ASPI) | • Condom use<br>• Safer sex intention | – *(only for women)*<br>– *(only for women)* |
| Pedersen & Blekesaune 2003 | 2101 | Norway | Range 20-25 | Stepwise multiple regression | 4-item scale | • Lifetime number of intercourse partners<br>• Sexual debut age | +<br>○ |
| Raj & Pollack 1995 | 103 | USA | M = 19.3 | Stepwise multiple regression | Index of Sexual Satisfaction (ISS) | • Contraceptive use | – |
| Rudolph et al 2020 | 1534 | USA | M = 18.3 (College women only) | Latent profile analysis | Sexual Satisfaction Scale for Women (SSS-W) | • Sexual Risk Survey (12 items), Risk profiles | – |
| Zimmer-Gembeck et al 2015 | 363 | Australia | M = 21.2 (Women only) | Multivariate regression | 1-item scale (sexual satisfaction) | • Number of sexual partners in previous 6 months<br>• Negative influence of alcohol/other drug use<br>• Condom use consistency | ○<br>–<br>○ |

N = sample size, + = positive correlation between risk behavior and sexual pleasure, – = negative correlation, ○ = no correlation, USA = United States of America, UK = United Kingdom, M = mean.

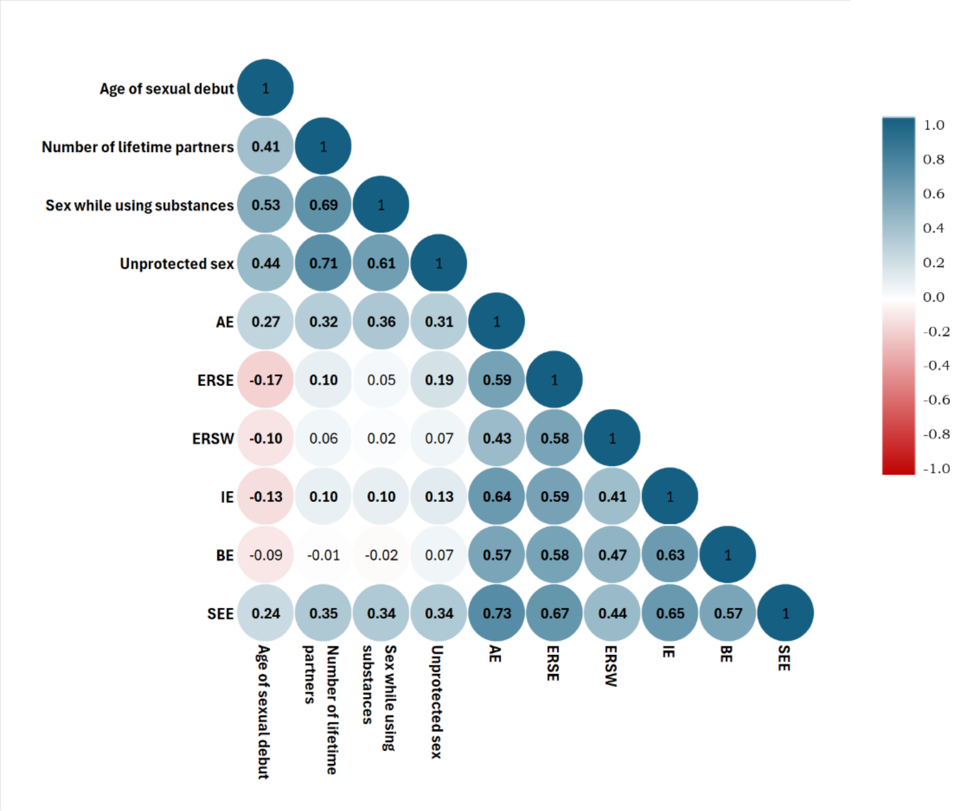


**Supplementary Figure S1.** Spearman's correlation matrix between Sexual Risk Behaviors and Sexual Pleasure subscales. AE = Arousal Enjoyment. ERSE = Enjoyment-Related Self-Efficacy. ERSW = Enjoyment-Related Self-Worth. IE = Interaction Enjoyment. BE = Bonding Enjoyment. SEE = Sexual Experience Enjoyment. Each value corresponds to the Spearman r value. $p<0.05$ (bold).

Supplementary Figure S1 shows the Spearman's correlation coefficients for the sexual pleasure subscales and SRBs. All correlations between SRBs were positive and significant. The same was true for the sexual pleasure subscales. Spearman's correlation coefficients for correlations between the sexual pleasure subscales and SRBs differed. Most correlations were positive and significant, while the age of sexual debut had negative correlations with four ASPI subscales (ERSE, ERSW, IE, BE). For Bonding Enjoyment, no correlations with SRBs were significant.
